# Developing a Core Outcome Set for capturing and measuring nurse wellbeing: A Delphi study

**DOI:** 10.1101/2024.10.14.24315438

**Authors:** Naomi Klepacz, David S. Baldwin, Gemma Simons

## Abstract

**Background:** Poor nurse wellbeing is a significant concern, adversely affecting patient care quality and satisfaction, contributing to poor job satisfaction, increased sickness absence and workforce retention issues. There are calls for evidence-based policies and interventions to address poor nurse wellbeing, but no consensus exists on how it should be captured and measured. We used a salutogenic and consensus approach to develop a core outcome set (COS) for capturing and measuring nurse wellbeing.

**Methods:** A Delphi methodology was employed. Participants were recruited from two stakeholder groups: 1) nurse wellbeing professionals, identified through relevant publications, conference/meeting attendance lists, and peer recommendations, and 2) Registered Nurses, recruited via social media, professional nursing bodies, and practitioner networks. The stakeholder panel completed two rounds of an online Delphi survey, rating 43 previously identified wellbeing outcomes on a nine-point Likert Scale, from ‘not important’ to ‘critical’. Consensus was defined as ≥75% of stakeholders agreeing a wellbeing outcome was critical for inclusion in the COS.

**Results:** Fifty-four stakeholders completed the first Delphi Round, and 45 participated in both rounds. Thirteen wellbeing outcomes met the *a-priori* threshold for inclusion in the COS: General Wellbeing, Health, Sleep, Positive Relationships, Personal Safety, Psychological Needs Satisfaction, Psychological Safety, Job Satisfaction, Morale, Life Work Balance, Compassion Satisfaction, Satisfaction with Patient Care, and Good Nursing Practice. The final COS was agreed by the stakeholder panel, without amendments.

**Conclusion:** This study establishes a COS for capturing and measuring nurse wellbeing. Implementing this COS has the potential to enable consistent data collection and evidence synthesis needed to support the development of nurse wellbeing strategies, policies and interventions. Future research will focus on identifying valid and reliable measurement tools.

**Trial Registration:** This study was prospectively registered with the COMET initiative www.comet-initiative.org (Registration: 2433)

## INTRODUCTION

Nurse wellbeing is an important indicator of the state of the nursing workforce. Poor nurse wellbeing impacts patient satisfaction and care quality, sickness absence, job satisfaction and leads to staff leaving the workforce.(1, 2, 3, 4, 5, 6) Wellbeing can be considered a continuum ranging from poor on one end to happiness, thriving and flourishing on the other. (7) However, most studies with nurses have focused on burnout (e.g.,(8, 9, 10)) and psychiatric morbidity (e.g.,(11, 12)), so little is known about positive (‘*salutogenic*’) indicators of wellbeing in this profession. (7)

Nurses constitute the largest group of clinical staff in the NHS, accounting for approximately 50% of the workforce. (13) Despite their deep commitment to providing high-quality patient care, many nurses experience poor wellbeing, stress and burnout. (7, 14) Physical and mental ill-health, burnout and exhaustion currently follow retirement as the top reason nurses leave the profession.(15) The 2023 NHS staff survey reports that 42% of nurses found their work emotionally exhausting, 46% experienced work-related stress, and 58% came to work despite not feeling well enough to perform their duties (so-called ‘presenteeism’).(16) While nurses strive to prevent their own sub-optimal wellbeing from adversely affecting patient care, (7) employers must recognise the direct link between nurse wellbeing and patient safety and satisfaction.(17) Ensuring nurse wellbeing is not only good for the nurses themselves but is essential for the health and safety of patients and key to nurse retention.

Nurse wellbeing is more than the absence of work-related stress, injury or disease; it is achieving good physical and mental health amongst the nursing workforce. (18) Nurse leaders have a professional responsibility to create healthy working environments that promote and sustain wellbeing. Managers, therefore, need a greater understanding of how nursing and the workplace impact nurse wellbeing and how to engage with staff who need support. (18) Effective decisions and strategies to improve nurse wellbeing must be grounded in reliable data, ensuring a robust evidence base. (19) A sharper focus on the drivers of positive nurse wellbeing is necessary to inform the development of policies, strategies, and interventions that will enable the nursing profession to flourish and thrive. However, wellbeing is a complex construct that includes measures and manifestations that have not yet been tested empirically among nurses. (7) No single measure can provide a complete picture of nurse wellbeing.

A Core Outcome Set (COS) offers an agreed minimum for what should be captured, measured and reported. (20) Our study takes a salutogenic (21) and consensus approach to developing a COS to capture and measure the wellbeing of nurses working in the NHS. It is anticipated that the consistent capture, measurement, and reporting of these outcomes will facilitate comparison by enhancing the ability to aggregate and analyse nurse wellbeing data, which is necessary to support policy, strategy and intervention development. This work builds on our previous study to develop a Core Outcome Set for capturing and measuring the wellbeing of doctors working in the NHS(22), so we have the additional objective of identifying potential convergence between the consensus outcomes for doctor and nurse wellbeing.

## METHODS

### Design

The study protocol was developed following Core Outcome Measures in Effectiveness Trials (COMET) criteria(23) and Core Outcome Set-STAndards for Development recommendations (COS-STAD)(24). It replicates our previous study, on doctor well-being.(22) The study was prospectively registered with the COMET Initiative(20) (Registration: 2433), and the findings are reported according to the Core Outcome Set-Standards for Reporting (COS-STAR) guidance.(25)

### Stakeholder Recruitment

A purposive sampling strategy was adopted to recruit a participant panel from two stakeholder groups: (i) Academics, policymakers, governance and support services staff, known here collectively as nurse wellbeing experts and (ii) Registered Nurses working in the NHS, considered experts by experience. Some overlap between the groups was anticipated, so participants were asked to self-assign to a stakeholder group based on their primary job role. The inclusion criterion for the nurse wellbeing experts group were: Individuals who have been or are involved in the concept, design, organisation, delivery, teaching, audit, governance, policy, guidance, research, or wellbeing of health and care professionals. We identified nurse wellbeing experts from relevant healthcare workforce wellbeing conferences, publications, and special interest groups by searching previous conference proceedings, published guidelines, and the wellbeing literature. We further identified these stakeholders through recommendations from others. Potential participants were emailed a study invitation. All registered nurses working in the NHS were eligible to participate; an invitation was disseminated through our research, clinical academic and practitioner networks, social media, nursing professional bodies and nursing Trade Unions. Invitations included links to the participant information sheet, a brief video outlining the study, and the online Delphi Survey. Participants were required to complete a consent form – the first page of the online Delphi survey - before registering their details (name and email) and indicating which of the two stakeholder groups they identified with. Demographic data, including age, gender, geographical location, clinical specialty (for nurses), ethnicity, and religion, were collected at registration. Each participant was assigned a Study ID at registration, ensuring data were anonymous at the point of collection.

### Outcomes and Domains

The starting point for this study was the list of 43 wellbeing outcomes used previously in the development of the Core Outcome Set for Doctor Wellbeing.(22) Using this set of outcomes allowed us to identify potential convergence between the consensus outcomes for doctor and nurse wellbeing. The 43 wellbeing outcomes are categorised into five domains: i) Overall appraisal of wellbeing, ii) Functional components of wellbeing, iii) Activity and participation components of wellbeing, iv) Work-related wellbeing, and v) Health and Care-specific Wellbeing.(26) The plain English descriptions of each outcome were reviewed for face validity, understanding and acceptability in a nursing context by our study advisory group (n=6) and modified according to feedback (Table 1).

**Table 1.** Outcomes and their descriptions by domain.

| Domain | Outcome | Description |
| --- | --- | --- |
| Overall appraisal of wellbeing | General Wellbeing | A state of positive feelings/affect/happiness and meeting full potential in the world (being the best person you can be in society). It can be measured subjectively and objectively using a salutogenic (positive) approach. |
|  | Meaning in life | Separate concept to wellbeing, subjective sense of purpose, engagement with a philosophy of life, or life-goals, and fulfilment. |
|  | Life satisfaction | Separate concept to wellbeing, subjective appraisal of how much the person likes the life they lead; one of the indicators of quality of life. |
|  | Quality of life | Separate concept to wellbeing, subjective appraisal of individuals position in life in the context of the culture and value system in which they live and in relation to their goals, expectations, standards, and concerns. |
|  | Wellness | Separate concept to wellbeing, subjective or objective evaluation of the active pursuit of behaviours, choices and lifestyles that lead to a state of holistic health. |
| Functional component of wellbeing | Vitality | Relaxed possession of energy (physical, mental, and emotional) and vigour; it is not actively strived for. |
|  | Optimism | Hopeful transcendence beyond (rising above) immediate circumstances. |
|  | Personality | Observable enduring characteristics/dispositions/tendencies to engage in certain patterns of behaviour. |
|  | Health | Subjective, or objective, evaluation of state of complete physical, mental, and social wellbeing, not merely the absence of disease or infirmity (the beneficial effects of green spaces, ability to relax, for example). |
|  | Physiological function | Objective (snapshot) of body functions i.e., Electroencephalography (EEG), Heart Rate Variability, Electrodermal activity (temperature, sweating, cortisol levels). |
|  | Cognitive function | Objective evaluation of domains such as, but not limited to, Attention, Memory, and Processing speed. |
|  | Self-esteem | Self-acceptance, self-worth (pride), sense of coherence (ability to predict events, belief in ability to manage them, that it is worth the effort, ability to be their true self, confidence in other achievements in non-work-related activities). |
|  | Sleep | Subjective, or objective, evaluation of duration, quality, and sense of feeling restored. |
|  | Financial security | Objective ability to pay for satisfactory accommodation, bills, care of dependents, ability to save for retirement, ability to cope with a sudden fall in income, ability to pay unexpected but necessary expenses. |
| Activity and participation component of wellbeing | Novelty | Subjective, or objective, growth through new experiences, learning (including post traumatic growth). |
|  | Positive relationships | Subjective, or objective, assessment of beneficial human connections (family and friends). |
|  | Sexual wellbeing | Subjective, or objective, assessment of sense of self and body, appreciating feelings of pleasure and desire, developing, and maintaining mutually respectful gender equal relationships, safe and pleasurable sexual interactions. |
|  | Recreational activity | Subjective, or objective, evaluation of the ability to participate and participation in non-work/leisure activities and the qualities of those chosen activities (example determinants are local investment and environment). |
|  | Diet | Subjective, or objective, evaluation of the nutritional content, quantity, and timing. |
|  | Physical activity | Subjective, or objective, assessment of the ability to participate in physical activity and the quality and quantity of physical exercise. |
|  | Engagement with preventative medicine | Subjective, or objective, assessment of participation in screening programmes they are eligible for and vaccines, accessing timely treatment. |
| Work-related wellbeing | Financial reward satisfaction | Subjective, or objective, evaluation of ability to receive gratification from financial reward for effort (for example satisfaction with pay and pension). |
|  | Personal safety | Subjective, or objective, ability to go about work, and get to and from work, free from threat and safe from physical or psychological harm (infection, radiation, bullying, theft, assault). |
|  | Psychological need satisfaction | Subjective, or objective, assessment of how autonomy (being in control of your life, work) belonging and competence needs have been supported by colleagues (inclusive, positive culture), managers (adequate workforce allow development), supporting services (IT, administration, legal, occupational health). |
|  | Psychological safety | Subjective, or objective, evaluation of the consequences of taking interpersonal risk at work (trust, information sharing). |
|  | Job satisfaction | Subjective, or objective, evaluation of how much they like their choice of work profession, specialism, roles. |
|  | Morale | Subjective, or objective, evaluation of feelings about the future, ability of an individual, group or organisation to have and meet shared goals/values. |
|  | Engagement | Subjective, or objective, assessment of involvement and absorption with, commitment to, work. |
|  | Life work balance | Subjective, or objective, quantity, quality, and equality of time away from work and at work, the salience/clarity of the roles (the ability to work flexibly). |
|  | Workability | Timely, objective assessment of having occupational competence and virtues, the health required for competence in an appropriate work environment by appropriate occupational health professionals. |
|  | Self-care | Subjective, or objective, assessment of behaviours to look after own health and wellbeing at work (taking breaks, time off work for sickness), accessing appropriate support services, adequate resources (estates, workforce, rapid-access self-referral services) to support this. |
|  | Professional Development | Subjective, or objective, assessment of ability to participate and engage with learning and teaching knowledge and skills, and to progress. |
|  | Identification with work | Subjective, or objective, assessment of value and meaning assumed by the individual, or a group/team, at work (pride in work, professional identity). |
|  | Resilience | Subjective, or objective, individual, or group level, preservation of, or return to, previous function after exposure to trauma. |
|  | Emotional intelligence | Subjective, or objective, self-awareness, self-management, social awareness, and relationship management. |
|  | Voice and influence | Subjective, or objective, assessment of ideas, concerns and expectations expressed informing policy and practice. |
|  | Confidence in leadership | Subjective or objective assessment of government and management competence, transparency and compassion, inclusivity, engagement and empowerment of those they are responsible and accountable for. |
|  | Recognition satisfaction | Subjective, or objective, evaluation of appreciation by colleagues, patients, public, government (civility). |
| Health and care specific wellbeing | Compassion satisfaction | Subjective evaluation of ability to receive gratification from caregiving to patients, patients' families, colleagues (satisfaction with non-financial rewards of the work). |
|  | Altruism | Subjective, or objective, evaluation of selfless concern for the wellbeing of others (patients and colleagues). |
|  | Satisfaction with patient care | Subjective, or objective, assessment of quality of health and social care their patients receive from themselves and others (impacted by things such as staffing levels, competence, equipment, estates and funding available). |
|  | Job plan/rota/rotation satisfaction | Subjective, or objective, evaluation of ability of role, responsibilities/rota/breaks to account for the quantity, types, of work (workload), the intensity, duration, of physical, mental, and emotional demands and the rest/activities/resources needed to maintain it. |
|  | Good nursing practice | Subjective or objective assessment of ability to engage with complex or challenging patients/cases and advocate for them as indicated and in an evidence-based way. |

### Delphi Survey and Analysis

The Delphi technique aims to generate consensus by collecting opinions from stakeholder panel members and is widely used in developing core outcome sets.(27) Using the online survey platform DelphiManager, (28) we listed the 43 wellbeing outcomes with plain English descriptions by domain. These were displayed in random order to participants. The Delphi survey was conducted over two rounds (Round 1 ran from 1 March 2023 to 24 March 2023, and Round 2 ran from 27 March to 30 April 2023). Adhering to the predefined Delphi survey guidelines,(23) we asked participants to rate the importance of including each outcome in the COS using a 9-point Likert Scale. For analysis, ratings were grouped: a rating of 1-3 on the Likert scale indicates the outcome is of ‘limited importance’ to include in the COS, a rating of 4-6 indicates the outcome is ‘important, but not critical’ to include, and a rating of 7-9 indicates that the outcome is ‘critical’ to include in a COS for the capture and measuring of nurses’ wellbeing. These groupings were devised by the Grading of Recommendations Assessment, Development and Evaluation (GRADE) working group and have been used widely for Delphi methods.(29) Participants had the opportunity to provide a rationale for their ratings, and were also given the option to indicate if they felt unable to score an outcome. At the end of each Delphi Round, participants had the opportunity to suggest additional outcomes that they felt were not included among the 43 wellbeing outcomes. Participants were advised that suggested outcomes should not be a symptom, sign or disease, nor a determinant of wellbeing. The criterion for including suggested outcomes in the next Delphi round was that the published definition of the outcome differed significantly from the plain English descriptions of the existing outcomes. Participants who suggested an additional outcome were emailed by the research team, with the justification for including or excluding the outcome based on this criterion, and offering participants the opportunity to present further evidence or explanation.

In Round 2, the percentage of stakeholder panel members giving each rating for an outcome was fed back to participants. Summary scores were not provided by stakeholder group as the opinions of both were equally important to the final COS. Participants were also reminded of their own ratings from Round 1 and were given the opportunity to revise their ratings after reviewing the feedback. Three email reminders were sent to participants to encourage the completion of a round.

The criteria for outcomes to be included in the COS were defined *a priori* as ≥75% of all participants rating an outcome as ‘critical for inclusion’ (rating 7-9). This aligns with our previous study(22) and other similar Core Outcome Sets (e.g., (30, 31, 32, 33)). The wellbeing outcomes that met this threshold for inclusion in the COS were communicated to all stakeholder panel members via email, along with an invitation to provide further comments and/or endorse the final COS.

### Ethical Approval

This study, which involved human participants, received approval from the University of Southampton Faculty Ethics Committee (ERGO 78343). Informed consent was obtained from all participants prior to their participation in this study.

## RESULTS

Study invitations were sent to 172 nurse wellbeing experts; of whom 33 consented and registered to participate, yielding a response rate of 19.2%. In addition, 29 Registered Nurses also agreed to participate, giving a total sample of 62 stakeholder panel participants. The mean age of participants was 48.1 years (range: 27 – 66 years); 48 participants (77.4%) self-identified as female, and 50 (80.7%) as White British. Four participants did not complete the survey (i.e. withdrew), and an additional four partially completed the survey. In total, 54 participants (87.1%) rated all 43 wellbeing outcomes (24 Registered Nurses, 30 nurse wellbeing experts). Participants who rated some or all of the outcomes in Round 1 (n=58) were invited to participate in Round 2, with 45 participants (18 Registered Nurses, 27 nurse wellbeing experts) completing Round 2, giving a retention rate from Round 1 to Round 2 of 87%. All participants in Round 2 rated all outcomes.

In Round 1, six participants submitted eight suggestions for additional outcomes (Supplementary Materials 1). None of these suggestions met the criteria to be included for consideration in Round 2, either because the definition of an existing outcome already captured them or because they were pathologies (for example, two participants suggested burnout for inclusion). However, based on these recommendations and participant feedback, the definition of ‘Identification with work’ was amended to include ‘professional identity’, and the definitions of ‘self-esteem’ and ‘identification with work’ were amended to include ‘pride’. None of the 43 outcomes were removed following Round 1. No additional outcomes were suggested in Round 2.

At the end of Round 2, 13 outcomes met the ≥75% threshold for inclusion in the Core Outcome Set for capturing and measuring nurse wellbeing. These outcomes were: General wellbeing, Health, Sleep, Positive Relationships, Personal safety, Psychological need satisfaction, Psychological safety, job satisfaction, Morale, Life work balance, Compassion satisfaction, Satisfaction with patient care, Good nursing practice (Table 2). These outcomes were subsequently emailed to participants for further comment and review. Participants agreed to the final COS without further amendments.

**Table 2.**
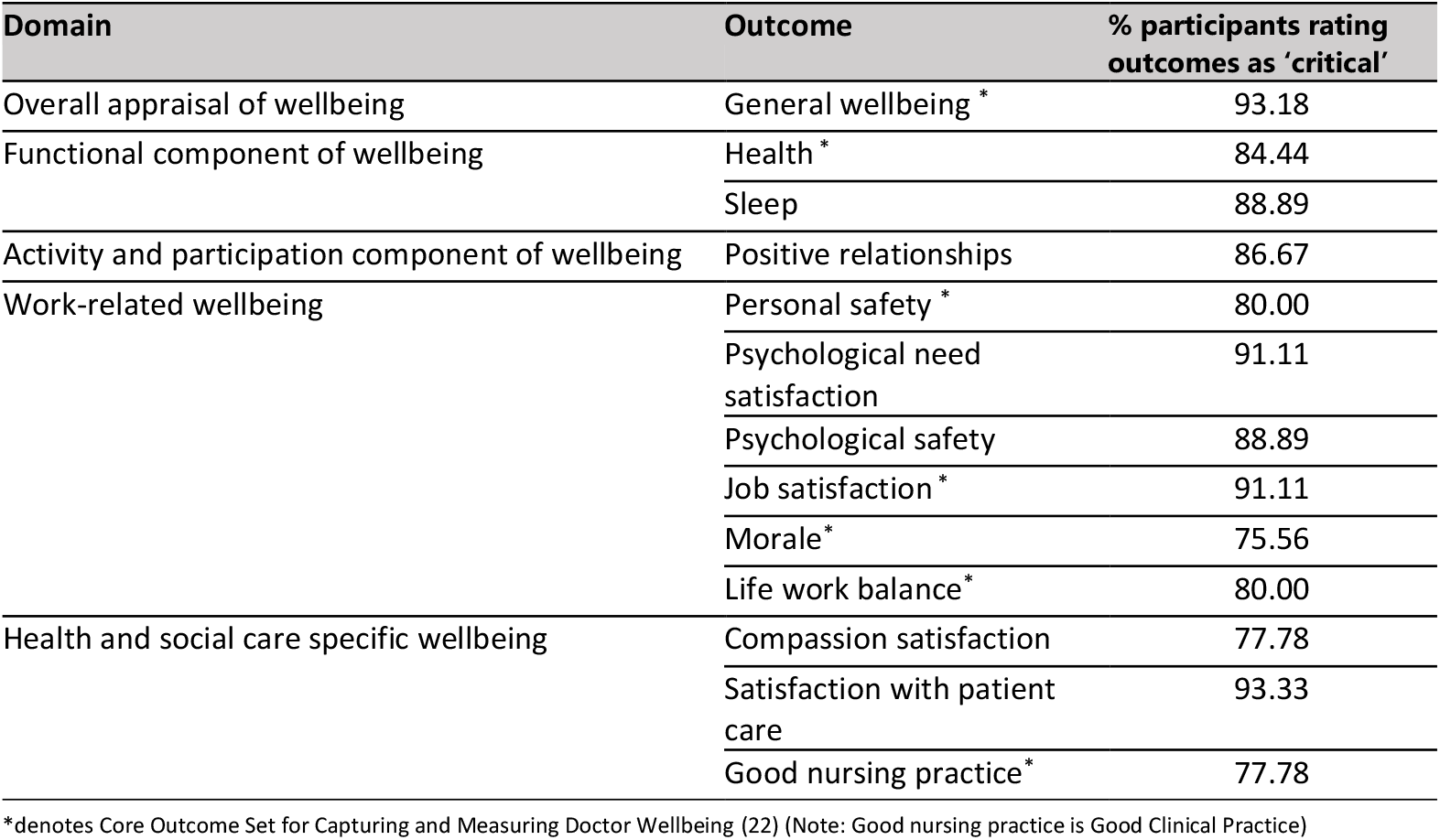
Final Core Outcome Set.

| Domain | Outcome | % participants rating outcomes as 'critical' |
| --- | --- | --- |
| Overall appraisal of wellbeing | General wellbeing * | 93.18 |
| Functional component of wellbeing | Health * | 84.44 |
|  | Sleep | 88.89 |
| Activity and participation component of wellbeing | Positive relationships | 86.67 |
| Work-related wellbeing | Personal safety * | 80.00 |
|  | Psychological need satisfaction | 91.11 |
|  | Psychological safety | 88.89 |
|  | Job satisfaction * | 91.11 |
|  | Morale* | 75.56 |
|  | Life work balance* | 80.00 |
| Health and social care specific wellbeing | Compassion satisfaction | 77.78 |
|  | Satisfaction with patient care | 93.33 |
|  | Good nursing practice* | 77.78 |

\*denotes Core Outcome Set for Capturing and Measuring Doctor Wellbeing (22) (Note: Good nursing practice is Good Clinical Practice)

## DISCUSSION

This study developed the first Core Outcome Set (COS) specifically for capturing and measuring the wellbeing of nurses working in the NHS. The stakeholder panel of registered nurses and nurse wellbeing experts reached a consensus on a minimum set of 13 outcomes that should routinely be captured and measured for nurse wellbeing. We recommend that future data collection initiatives adopt this COS to ensure standardisation, enabling a consistent, comparable, and comprehensive evidence-base with the potential to support decision-making for policy and practice. No prior COS has been developed for the wellbeing of nurses. Previous research has focused on determinants and interventions of nurse wellbeing rather than the outcomes that might demonstrate how these determinants or interventions influence this profession. By creating this COS and promoting its use, we seek to shift the current discourse towards an understanding of positive (salutogenic) components of nurse wellbeing. This shift is critical for the development of effective wellbeing policies, strategies and interventions that empower nurses to flourish in the workplace. Future research is now needed to identify and evaluate outcome measurement instruments.

A strength of our approach is that it provides outcomes for each of the five wellbeing domains. Several agreed-upon wellbeing outcomes, such as morale, personal safety, and job satisfaction, are already captured through, for example, the NHS staff survey (34) and the RCN Employment survey.(35) Whereas other outcomes, such as ‘good nursing practice,’ will require the identification of outcome measurement instruments based on their descriptions in Table 1. Furthermore, the seven wellbeing outcomes that comprise the COS for capturing and measuring doctor’s wellbeing (General wellbeing, Health, Personal Safety, Job satisfaction, Morale, Life-work balance, and Good Clinical Practice)(22) met the threshold for inclusion in the COS for nurses’ wellbeing. This alignment suggests that factors considered relevant to doctors’ wellbeing are similarly relevant to nurses. Indeed, previous research indicates that certain features of work-related wellbeing and mental ill-health are common across all NHS staff groups.(6) However, the additional outcomes identified for the COS for nurse wellbeing underscore important profession-specific differences that must be considered when developing policies, strategies, and interventions for nurses.

The methodology used in this study was robust and replicable, following the COS-STAD guidelines (24) and built on our previous work developing a COS for capturing and measuring doctor wellbeing. (22) The long list of outcomes presented to the stakeholder panel was evidence-based, (26) and our study advisory group ensured the relevance and validity of this list to nursing. The presentation of domains to the stakeholder panel participants was randomised using the DelphiManager platform (28) to avoid presentation bias. Furthermore, additional wellbeing outcomes suggested by participants during the Delphi survey were already represented by existing wellbeing outcomes, further supporting the comprehensiveness of the long list. The suggestions to add burnout as a wellbeing outcome reflect the current use of burnout and psychiatric morbidities as proxies for wellbeing, further underscoring the need for this COS. While the lack of an internationally agreed-upon operational definition of nurse wellbeing may be seen as a limitation, we addressed this by utilising our published operational definition of wellbeing (21) and the application of a salutogenic, consensus-based methodology. This approach enabled us to establish a panel-agreed COS for wellbeing outcomes relevant to nurses.

A further strength of this study was that it included registered nurses and nurse wellbeing professionals in the stakeholder panel. However, we acknowledge that stakeholders outside the present panel might have differing views. The sample size was appropriate for a Delphi study(24), as was the response rate from nurse wellbeing professionals to invitation and the overall retention rate. Our focus on nurses working in the UK’s National Health Service (NHS) means that stakeholders were invited accordingly. The recruitment method for registered nurse stakeholders was designed to reach all nurses working in the NHS; however, we are unaware of how many potential participants saw our invitation and elected not to participate. While this COS might be relevant to nurses working in other healthcare systems – both in the UK and beyond – additional investigation is required to ensure its broader applicability.

We acknowledge that users of this COS may find it challenging to capture and measure all 13 outcomes that comprise this COS, and it should be noted that the feasibility of using this COS on every occasion nurse wellbeing is measured has not yet been tested. While we suggest these outcomes as a minimum, users may wish to include other outcomes relevant to their research or capture and measure only those outcomes from their domain of interest; for example, the work-related wellbeing domain with its six wellbeing outcomes or the subset of seven wellbeing outcomes common to both doctors and nurses. The robust methodology we have applied in this study could be repeated to assess the relevance of these outcomes to other healthcare professions. This COS provides a framework to better understand positive components of wellbeing in the nursing profession, and in line with COMET guidelines (23), our next step is to identify which outcome measurement instruments would be most appropriate and accessible for end users.

## CONCLUSION

This study has identified a minimum set of wellbeing outcomes that should be used when measuring NHS nurse wellbeing. Implementing this COS will reduce heterogeneity in measurement approaches, facilitating evidence synthesis and benchmarking to better understand the current state of nurse wellbeing. Future efforts will focus on identifying and evaluating the most appropriate instruments for measuring these outcomes.

## Author Contributions

- GS and DSB devised the study and acquired funding.
- NK designed the study, collected, analysed, and interpreted data, wrote the manuscript, and edited and approved the final article.
- GS and DSB supervised the study’s design, data collection, analysis, and interpretation and approved the final manuscript.
- GS acts as guarantor.

## Disclaimer

The views expressed in this publication are those of the authors and not necessarily those of the National Institute for Health and Care Research or the Department of Health and Social Care.

## Acknowledgements

The authors would like to thank all those who participated in the Delphi survey and provided feedback on all stages of the research process, NIHR ARC Wessex for funding this research, Catherine Smith and Jane Ball for advising on the early stages of this study, and Professor Peter Griffiths for reviewing the draft manuscript.

## Data availability statement

All data relevant to the study are included in the article or uploaded as supplementary materials.

## Ethics statement

This study involved human participants and was approved by the University of Southampton Faculty Ethics Committee (ERGO 78343). Participants gave informed consent before participating.

## REFERENCES

1. Sizmur S, Raleigh V. The risks to care quality and staff wellbeing of an NHS system under pressure. Oxford: King’s Fund; 2018.

2. Aiken L, Sloane DM, Ball J, Bruyneel L, Rafferty AM, Griffiths P. Patient satisfaction with hospital care and nurses in England: an observational study. BMJ open. 2021;8.

3. Maben J, Adams M, Peccei R, Murrells T, Robert G. ‘Poppets and parcels’: the links between staff experience of work and acutely ill older peoples’ experience of hospital care. Int J Older People Nurs. 2012;7(2):83–94.

4. Hall LH, Johnson J, Watt I, Tsipa A, O’Connor DB. Healthcare Staff Wellbeing, Burnout, and Patient Safety: A Systematic Review. PLoS One. 2016;11(7):e0159015.

5. Boorman S. NHS Health and Well-being Review: Interim Report and Final Report. London; 2009.

6. Taylor C, Maben J, Jagosh J, Carrieri D, Briscoe S, Klepacz N, et al. Care Under Pressure 2: a realist synthesis of causes and interventions to mitigate psychological ill health in nurses, midwives and paramedics. BMJ Qual Saf. 2024:bmjqs-2023-016468.

7. Kinman G, Teoh K, Harriss A. The Mental Health and Wellbeing of Nurses and Midwives in the United Kingdom. London: The Society of Occupational Medicine; 2020.

8. Van der Heijden B, Brown Mahoney C, Xu Y. Impact of Job Demands and Resources on Nurses’ Burnout and Occupational Turnover Intention Towards an Age-Moderated Mediation Model for the Nursing Profession. IJERPH. 2019;16(11):2011.

9. Khatatbeh H, Pakai A, Al-Dwaikat T, Onchonga D, Amer F, Premusz V, et al. Nurses’ burnout and quality of life: A systematic review and critical analysis of measures used. Nurs Open. 2022;9(3):1564–74.

10. Shah MK, Gandrakota N, Cimiotti JP, Ghose N, Moore M, Ali MK. Prevalence of and Factors Associated With Nurse Burnout in the US. JAMA Network Open. 2021;4(2):e2036469–e.

11. Barleycorn D. Awareness of secondary traumatic stress in emergency nursing. Emergency Nurse. 2019;27(5):19–22.

12. Slusarska B, Nowicki GJ, Niedorys-Karczmarczyk B, Chrzan-Rodak A. Prevalence of Depression and Anxiety in Nurses during the First Eleven Months of the COVID-19 Pandemic: A Systematic Review and Meta-Analysis. IJERPH. 2022;19(3):1154.

13. NHS Digital. NHS Workforce Statistics, December 2023 England and Organisation 2023 [Available from: https://digital.nhs.uk/data-and-information/publications/statistical/nhs-workforce-statistics/december-2023.

14. Van Bogaert P, Wouters K, Willems R, Mondelaers M, Clarke S. Work engagement supports nurse workforce stability and quality of care: nursing team-level analysis in psychiatric hospitals. Journal of Psychiatric and Mental Health Nursing. 2013;20(8):679–86.

15. Nursing and Midwifery Council. 2023 NMC Register Leavers Survey. 2023.

16. NHS England. NHS Staff Survey Interactive Dashboard (Results of 2023 survey) 2024 [Available from: https://nhssurveys.co.uk/nss/survey-information/.

17. Maben J, Peccei R, Adams M, Robert G, Richardson A, Murrells T, et al. Exploring the relationship between patients’ experiences of care and the influence of staff motivation, affect and wellbeing. 2012.

18. Royal College of Nursing. Nursing Workforce Standards. London: Royal College of Nursing; 2021 07/05/2021. Contract No.: 009 681.

19. Bernardi FA, Alves D, Crepaldi N, Yamada DB, Lima VC, Rijo R. Data Quality in Health Research: Integrative Literature Review. J Med Internet Res. 2023;25:e41446.

20. COMET Initiative. Core Outcome Measures in Effectiveness Trials (COMET Initiative) database n.d [Available from: https://www.comet-initiative.org/.

21. Simons G, Baldwin DS. A critical review of the definition of ‘wellbeing’ for doctors and their patients in a post Covid-19 era. Int J Soc Psychiatry. 2021;67(8):984–91.

22. Simons G, Klepacz N, Baldwin DS. Which outcomes should be included in a core outcome set for capturing and measuring doctor well-being? A Delphi study. medRxiv. 2024:2024.04.11.24305668.

23. Williamson PR, Altman DG, Bagley H, Barnes KL, Blazeby JM, Brookes ST, et al. The COMET Handbook: version 1.0. Trials. 2017;18(Suppl 3):280.

24. Kirkham JJ, Davis K, Altman DG, Blazeby JM, Clarke M, Tunis S, et al. Core outcome Set-STAndards for Development: the COS-STAD recommendations. PLoS medicine. 2017;14(11):e1002447.

25. Kirkham JJ, Gorst S, Altman DG, Blazeby JM, Clarke M, Devane D, et al. Core Outcome Set-STAndards for Reporting: The COS-STAR Statement. PLoS Med. 2016;13(10):e1002148.

26. Simons G. How should wellbeing be measured in UK doctors? A salutogenic, consensus approach, towards a Core Outcome Set for doctor wellbeing measurement: University of Southampton; 2022.

27. International Consortium Health Outcome Measurement. Our mission n.d. [

28. COMET Initiative. DelphiManger 2023 [Available from: https://www.comet-initiative.org/delphimanager/.

29. Guyatt GH, Oxman AD, Vist GE, Kunz R, Falck-Ytter Y, Alonso-Coello P, et al. GRADE: an emerging consensus on rating quality of evidence and strength of recommendations. BMJ. 2008;336(7650):924–6.

30. Vogel C, Zwolinsky S, Griffiths C, Hobbs M, Henderson E, Wilkins E. A Delphi study to build consensus on the definition and use of big data in obesity research. Int J Obes (Lond). 2019;43(12):2573–86.

31. Knaapen M, Hall NJ, van der Lee JH, Butcher NJ, Offringa M, Van Heurn EWE, et al. Establishing a core outcome set for treatment of uncomplicated appendicitis in children: study protocol for an international Delphi survey. BMJ Open. 2019;9(5):e028861.

32. Santaguida P, Dolovich L, Oliver D, Lamarche L, Gilsing A, Griffith LE, et al. Protocol for a Delphi consensus exercise to identify a core set of criteria for selecting health related outcome measures (HROM) to be used in primary health care. BMC Fam Pract. 2018;19(1):152.

33. Blade J, Calleja MA, Lahuerta JJ, Poveda JL, de Paz HD, Lizan L. Defining a set of standardised outcome measures for newly diagnosed patients with multiple myeloma using the Delphi consensus method: the IMPORTA project. BMJ Open. 2018;8(2):e018850.

34. NHS Staff Survey Coordination Centre. Technical guide to the 2023 staff survey data (Version 2). 2023.

35. Royal College of Nursing. State of the Profession Report: RCN Employment Survey 2023. London; 2024 May 2024. Contract No.: 011 484.

