## Supplementary Materials 1 for "Developing a Core Outcome Set for capturing and measuring nurse wellbeing: A Delphi study"

**Supplementary Materials 1**. Additional outcomes suggested by stakeholders in Round 1 with rationale for not including as unique outcomes in Round 2.

|  | **Suggested New Outcomes** | **Rationale for not including as a unique outcome in Round 2** |
| --- | --- | --- |
| 1 | Sharing concerns with others | **Not novel, captured by existing outcomes** This is captured by the outcome ‘Psychological Safety’. |
| 2 | Supportive colleague interaction/relationships | **Not novel, captured by existing outcomes** This is captured by the outcome ‘Psychological Safety’ |
| 3 | Burnout | **Diagnosis or pathology** We chose not to include mental health diagnoses as wellbeing outcomes, as these are separate concepts. |
| 4 | Belonging | **Not novel, captured by existing outcomes** This is captured by the outcome ‘Psychological Needs Satisfaction’ |
| 5 | Human flourishing | **Not novel, captured by existing outcomes** This is captured by the outcomes ‘Vitality’ and ‘Novelty’ |
| 6 | Professional identity | **Not novel, captured by existing outcomes** This is captured by the outcome ‘identification with work’. |
| 7 | Pride (in being part of the organisation for which they nurse works) | **Not novel, captured by existing outcomes** This is captured by the outcomes ‘self-esteems’ and ‘identification’. |
| 8 | Burnout | **Diagnosis or pathology** We chose not to include mental health diagnoses as wellbeing outcomes, as these are separate concepts. |
