## Supplementary Materials 2 for "Developing a Core Outcome Set for capturing and measuring nurse wellbeing: A Delphi study"

| **Table 1**. Percentage agreement for outcomes at the end of Round 2. | | | | |
| --- | --- | --- | --- | --- |
| Domain | Outcome | Limited Importance  % | Important  % | Critical Importance  % |
| Overall appraisal of wellbeing | General wellbeing | 0.00 | 6.82 | 93.18* |
|  | Meaning in life | 6.67 | 22.22 | 71.11 |
|  | Life satisfaction | 0.00 | 37.78 | 62.22 |
|  | Wellness | 0.00 | 26.27 | 73.33 |
| Functional component of wellbeing | Vitality | 4.55 | 59.09 | 36.36 |
|  | Optimism | 0.00 | 68.89 | 31.11 |
|  | Personality | 33.33 | 54.76 | 11.90 |
|  | Health | 0.00 | 15.56 | 84.44* |
|  | Physiological function | 4.44 | 46.67 | 48.89 |
|  | Cognitive function | 2.22 | 42.22 | 55.56 |
|  | Self-esteem | 0.00 | 42.22 | 57.78 |
|  | Sleep | 2.22 | 8.89 | 88.89* |
|  | Financial security | 0.00 | 42.22 | 57.78 |
| Activity and participation component of wellbeing | Novelty | 27.91 | 69.77 | 2.33 |
|  | Positive relationships | 2.22 | 11.11 | 86.67* |
|  | Sexual wellbeing | 34.88 | 58.14 | 6.98 |
|  | Recreational activity | 4.55 | 77.27 | 18.18 |
|  | Diet | 4.55 | 56.82 | 38.64 |
|  | Physical activity | 4.55 | 38.64 | 56.82 |
|  | Engagement with preventative medicine | 9.09 | 63.64 | 27.27 |
| Work-related wellbeing | Financial reward satisfaction | 2.22 | 57.78 | 40.00 |
|  | Personal safety | 0.00 | 20.00 | 80.00* |
|  | Psychological need satisfaction | 0.00 | 8.89 | 91.11* |
|  | Psychological safety | 0.00 | 11.11 | 88.89* |
|  | Job satisfaction | 0.00 | 8.89 | 91.11* |
|  | Morale | 0.00 | 24.44 | 75.56* |
|  | Engagement | 0.00 | 42.22 | 57.78 |
|  | Life work balance | 0.00 | 20.00 | 80.00* |
|  | Workability | 2.38 | 57.14 | 40.48 |
|  | Self-care | 0.00 | 44.44 | 55.56 |
|  | Professional development | 4.44 | 33.33 | 62.22 |
|  | Identification with work | 2.27 | 54.55 | 43.18 |
|  | Resilience | 6.98 | 34.88 | 58.14 |
|  | Emotional intelligence | 11.11 | 46.67 | 42.22 |
|  | Voice and influence | 0.00 | 46.67 | 53.33 |
|  | Confidence in leadership | 0.00 | 31.11 | 68.89 |
|  | Recognition satisfaction | 0.00 | 46.67 | 53.33 |
|  | Compassion satisfaction | 0.00 | 22.22 | 77.78* |
| Health and social care specific wellbeing | Altruism | 8.89 | 64.44 | 26.67 |
|  | Satisfaction with patient care | 0.00 | 6.67 | 93.33* |
|  | Role/responsibility/rota/break satisfaction | 2.22 | 37.78 | 60.00 |
|  | Good nursing practice | 0.00 | 22.22 | 77.78* |

*Ratings that met the threshold (≥75%) to be included in the COS.
